# Validation of molecular clock inferred HIV infection ages: evidence for accurate estimation of infection dates

**DOI:** 10.1101/2020.12.11.20247601

**Authors:** E. G. Kostaki, S. Limnaios, S. Roussos, M. Psichogiou, G.K. Nikolopoulos, S-R. Friedman, A. Antoniadou, M. Chini, A. Hatzakis, V. Sypsa, G. Magiorkinis, C. Seguin-Devaux, D. Paraskevis

**Author notes:** **Correspondence to** Dimitrios Paraskevis, PhD, Associate Professor of Epidemiology and Preventive Medicine, Department of Hygiene, Epidemiology and Medical Statistics, Medical School, National and Kapodistrian University of Athens, 75 Mikras Asias Street, GR-11527, Athens, Greece.

## Abstract

Improving HIV diagnosis, access to care and effective antiretroviral treatment provides our global strategy to reduce HIV incidence. To reach this goal we need to increase our knowledge about local epidemics. HIV infection dates would be an important information towards this goal, but they are largely unknown. To date, methods to estimate the dates of HIV infection are based mainly on laboratory or molecular methods. Our aim was to validate molecular clock inferred infection dates that were estimated by analysing sequences from 145 people living with HIV (PLHIV) with known transmission dates (clinically estimated infection dates). All HIV sequences were obtained by Sanger sequencing and were previously found to belong to well-established molecular transmission clusters (MTCs). Our analysis showed that the molecular clock inferred infection dates were correlated with the clinically estimated ones (Spearman’s Correlation coefficient = 0.93, p<0.001) and that there was an agreement between them (Lin’s concordance correlation coefficient = 0.92, p<0.001). For most cases (61.4%), the molecular clock inferred preceded the clinically estimated infection dates. The median difference between clinically and molecularly estimated dates of infection was of 0.18 (IQR: -0.21, 0.89) years. The lowest differences were identified in people who inject drugs of our study population. Our study shows that the estimated time to more recent common ancestor (t_MRCA_) of nodes within clusters provides a reliable approximation of HIV infections for PLHIV infected within MTCs. Next-generation sequencing data and molecular clock estimates based on heterochronous sequences provide, probably, more reliable methods for inferring infection dates. However, since these data are not available in most of the HIV clinical laboratories, our approach, under specific conditions, can provide a reliable estimation of HIV infection dates and can be used for HIV public health interventions.

## Introduction

HIV infection is still expanding around the world. According to WHO/UNAIDS, the number of people living with HIV (PLHIV) globally was 38 million in 2019. In the absence of an effective vaccine, the goal to reduce HIV incidence is based on the 90-90-90 target for diagnosis, access to care and effective antiretroviral (ARV) treatment. Knowledge about local HIV epidemics would facilitate the implementation and success of this ambitious global target (1). One of the critical epidemiological parameters is HIV incidence, but HIV infection dates are largely unknown. This is due to the lack of seronegative tests for the majority of the PLHIV, as well as the rare diagnosis during the acute phase of infection.

To date, the available methods to estimate HIV infection are based on serological or genomic-based approaches. Laboratory (serological) methods, such as the Limiting Antigen Avidity Assay (LAg) or the Calypte Incidence Assay (BED), can classify recent from non-recent infections (2-4). Genomic-based or molecular methods can estimate the age of HIV infection based on previous observations that HIV genetic divergence increases over time. Several methods have been described so far including high resolution melting assays (HMA), the degree of sequence ambiguities in Sanger population sequencing (5-7), molecular clock-based methods (8, 9) and the Hamming distances between different quasispecies (7, 10). More recently, the utilization of next-generation sequencing (NGS), and specifically the deep NGS, provided a new advance for the estimation of the time since HIV infection (11-13).

Our aim was to investigate if time to the most recent common ancestor (t_MRCA_) of nodes within well-established molecular transmission clusters (MTCs) can be used as a proxy for the HIV infection date, using Sanger sequencing data from densely sampled populations with known transmission dates (clinically estimated infection dates).

## Materials and Methods

Our study population consisted of PLHIV with known transmission dates from the following datasets: i) a “seek-test-treat” intervention programme named “ARISTOTLE” implemented in Athens, Greece, during 2012-2013 (14), ii) a network-based intervention that targeted persons with recent HIV-infection (“TRIP”) implemented in Athens, Greece, between 2013 and 2015 (15), iii) PLHIV infected within a subtype A1 MTC in Greece (16, 17), iv) People who inject drugs (PWID) infected locally during an HIV outbreak in Luxembourg since 2014 and v) PLHIV infected within a CRF42_BF MTC in Luxembourg (18) (Table 1). HIV sequences were selected to belong to PWID-specific MTCs [datasets (i), (ii) and (iv)] and local MTCs [datasets (iii) and (v)] (19). Local MTCs were defined as phylogenetic clusters including ≥ 2 sequences from the same geographic area (Greece or Luxembourg) at a proportion greater than 70% compared to the total number of sequences within the cluster, as previously described in detail (16, 20). All sequences were generated before the ARV treatment initiation. Each patient was specified by a unique serial number to avoid double-counting. Duplicate patients were detected and excluded from the study population.

**Table 1.** Number and risk group of people living with HIV of the study population, sampling period, HIV-1 subtype and country of sampling of the sequences under study.

| **Characteristic** | **PLHIV**<br>**(n)** | **Risk**<br>**group** | **Sampling**<br>**period** | **HIV-1**<br>**subtype** | **Country of**<br>**sampling** |
| --- | --- | --- | --- | --- | --- |
| Overall | 145 |  | 2003 – 2018 |  |  |
| Dataset |  |  |  |  |  |
| i & ii | 36 | PWID | 2012 – 2013 |  | Greece |
| iii | 24 | Non-PWID | 2008 – 2014 | A1 | Greece |
| iv | 46 | PWID | 2005 – 2018 |  | Luxembourg |
| v | 39 | Non-PWID | 2003 – 2017 | CRF42_BF | Luxembourg |

PLHIV, people living with HIV; PWID, people who inject drugs

Known transmission dates were available from: a) the midpoint between the last available seronegative and the first seropositive tests for HIV, the possible date of HIV acquisition based on the diagnosis of acute HIV infection and patient self-report [datasets (i), (iii), (iv) and (v)] (17), or b) the putative date of HIV infection which was determined using the LAg method [dataset (ii)] (2, 15). From now on we will refer at these dates as “clinically estimated infection dates”.

Molecular clock calculation using phylodynamic analysis was used for the re-estimation of the infection dates of our study population (PLHIV with an available clinically estimated infection date). Phylodynamic analyses were applied on PR and partial RT sequences generated by Sanger sequencing and implemented in BEAST (versions 1.8.0 and 2.1.3), using different nucleotide substitution models (GTR+G, HKY+G), uncorrelated lognormal relaxed clock models and the birth-death models (21, 22). The full description of these analyses has been provided in detail elsewhere (17, 19, 23). The age of infection (infection dates) was approximated to the median estimate of the t_MRCA_ of the internal nodes within each MTC. We will refer at these dates as “molecular clock inferred infection dates”.

Medians, interquartile ranges (IQRs) and 95% confidence intervals (CIs) were used to summarize the data. Spearman’s correlation coefficient was used to measure the strength and direction of association between the clinically estimated and molecular clock inferred infection dates, while Lin’s concordance correlation coefficient was used to measure the agreement between these estimates. The fitted regression line was estimated by a quartile (median) regression model and was compared to the line of equality by testing the two-tailed hypothesis of slope = 1 and intercept = 0. In addition, the Bland-Altman plot was used to quantify and describe the agreement between the two variables and the presence of outliers. Spearman’s correlation coefficient was also used to assess putative correlation between the difference of the clinically and molecularly estimated dates of infection and the time interval from the last seronegative to the first seropositive tests for HIV, for a subset of our study population (26 PLHIV from whom the clinically estimated infection date was represented by the midpoint between these tests). The level of significance was set at 0.05. Analyses were performed in Stata 13-StataCorp LP software.

## Results

The total number of HIV sequences used in the context of our study was 145. These sequences were retrieved from five different datasets, as it was described in detail previously in the methods section. The samples from “ARISTOTLE” [dataset (i), n=23] and “TRIP” [dataset (ii), n=13] were merged since they were drawn from PWID participating in intervention programmes implemented in Athens, Greece, during the same time period (Table 1). The data from Luxembourg included both samples from PWID infected during an outbreak [dataset (iv), n=46] and non-PWID found within a CRF42_BF MTC expanding locally [dataset (v), n=39] (Table 1). The sampling periods spanned over a long time between 17 June 2003 and 11 January 2018 for the data from Luxembourg [datasets (iv) and (v)], and 22 May 2008 - 05 December 2014 for the subtype A1 data from Greece [dataset (iii), n=24] (Table 1).

The molecular clock inferred infection dates were positively correlated with the clinically estimated infection dates with the Spearman’s Correlation coefficient equal to 0.93 (p<0.001) (Figure 1). For most cases, the molecular clock inferred infection dates preceded the clinically estimated ones (89 of 145, 61.4%). Moreover, the fitted regression line was described by the equation: molecular clock inferred = −53.49 + 1.03 × clinically estimated, with 95% CI for the estimated intercept at (−138.49, 31.51) (p=0.216) and with 95% CI for the estimated slope at (0.98, 1.07) (p=0.0514) (Figure 1). Lin’s concordance correlation coefficient was 0.92 (95% CI: 0.90, 0.94) and showed an agreement between clinically and molecularly estimated infection dates (p<0.001).

**Figure 1.**
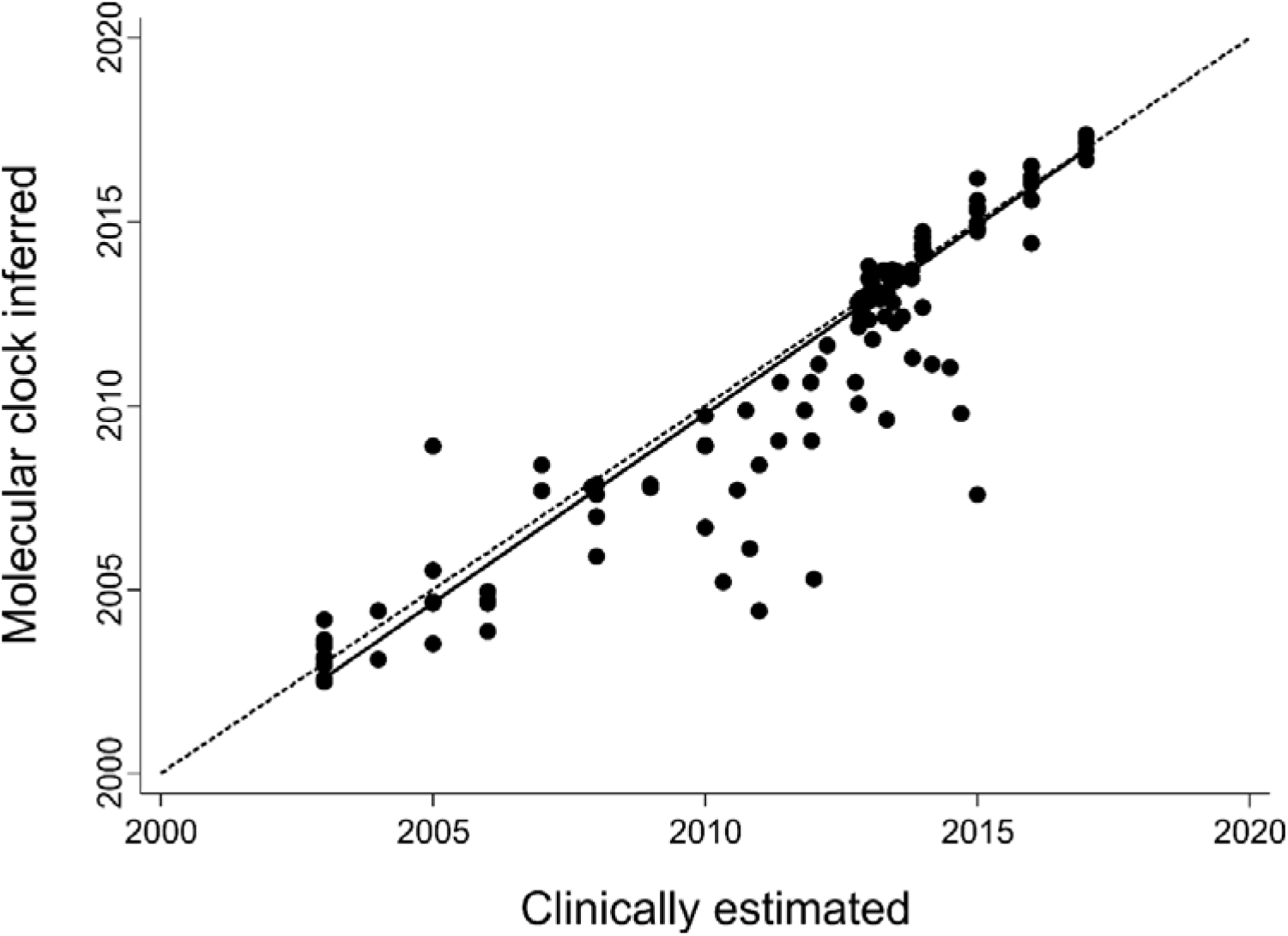
Scatter plot showing relationship between molecular clock inferred infection dates (shown along the y-axis) and clinically estimated infection dates (shown along the x-axis) for 145 people living with HIV of the study population. Each individual is represented by one circle. The solid line corresponds to the regression fitted line and the dashed line to the line of equality. Spearman’s correlation coefficient: r=0.93 (p<0.001).

The median difference between the clinically estimated and molecular clock inferred infection dates was 0.18 (IQR: −0.21, 0.89) years across all sequences (n=145) (Table 2). The largest differences were found for the non-PWID infected with subtype A1 in Greece [dataset (iii), median value: 2.04 (IQR: 0.62, 2.99) years] and for the non-PWID of the CRF42_BF clade in Luxembourg [dataset (v), median value: 0.52 (IQR: −0.16, 1.28) years] (Table 2). Generally, for the PWID of our study population the differences were very low [datasets (i) & (ii): median value: 0.09 (IQR: −0.04, 0.36) years; dataset (iv): median value: −0.24 (IQR: −0.46, 0.19) years) (Table 2). The differences between the clinically and molecularly estimated infection dates for all PLHIV of our study population were plotted against the mean obtained by the two variables (Figure 2). The Bland–Altman plot revealed an agreement between the two variables, an absence of a consistent bias and the existence of a small number of outliers [6 of 145 (4.1%) PLHIV with a difference greater than 4 years] (Figure 2). Moreover, the difference between the clinically estimated and molecular clock inferred infection dates was not correlated with the time interval from the last seronegative to the first seropositive tests (p=0.822).

**Table 2.** Difference between clinically estimated and molecular clock inferred infection dates and time interval between molecular clock inferred infection dates and sampling dates for the study population.

| Characteristic | Difference between<br>estimated infection dates,<br>in years, median (IQR) | Time interval between<br>infection and sampling,<br>in years, median (IQR) |
| --- | --- | --- |
| Overall | 0.18 (-0.21, 0.89) | 0.59 (0.25, 1.45) |
| Dataset |  |  |
| i & ii | 0.09 (-0.04, 0.36) | 0.32 (0.18, 0.60) |
| iii | 2.04 (0.62, 2.99) | 2.42 (1.09, 3.46) |
| iv | -0.24 (-0.46, 0.19) | 0.34 (0.14, 0.59) |
| v | 0.52 (-0.16, 1.28) | 1.26 (0.38, 2.13) |

IQR, interquartile range

**Figure 2.**
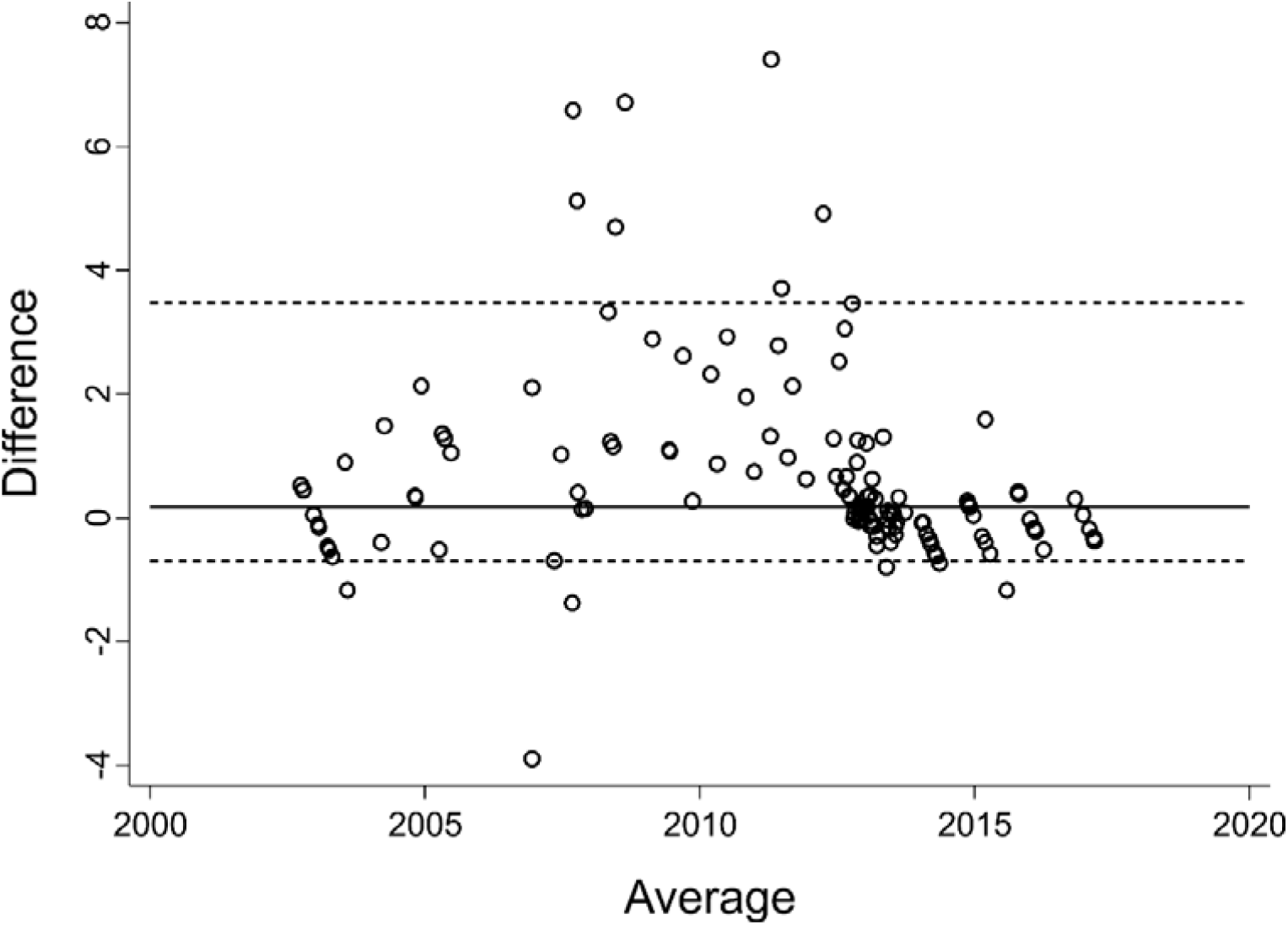
Bland-Altman plot showing the differences (in years) between the clinically estimated and molecular clock inferred infection dates (shown along the y-axis) compared with the mean of the dates (x-axis). The dashed lines represent the 5th and 95th percentiles for the differences and the solid line corresponds to the median difference.

The median estimate of the time interval between the molecular clock inferred infection dates and the sampling dates of the sequences was 0.59 (IQR: 0.25, 1.45) years (Table 2). As for the difference between the clinically and molecularly estimated infection dates, the largest values of this time interval were found for the non-PWID infected with subtype A1 [dataset (iii), median value: 2.42 (IQR: 1.09, 3.46) years] and CRF42_BF [dataset (v), median value: 1.26 (IQR: 0.38, 2.13) years] (Table 2). For PWID infected in Luxembourg [dataset (iv)] the median value of the interval was 0.34 (IQR: 0.14, 0.59) years, however this value was small and comparable to that for PWID infected in Athens, Greece [datasets (i) & (ii), median value: 0.32 (IQR: 0.18, 0.60)

## Discussion

Knowledge of infection dates is valuable for estimating the number of new HIV infections per year, the improvement of linkage to care and also for tailoring efforts to control HIV transmission (24, 25). Our study provides evidence that, under specific conditions, HIV data generated by Sanger sequencing can be used for reliable estimation of HIV infection dates. Specifically, the t_MRCA_ of nodes within local MTCs can provide a proxy of the HIV infection dates. Our validation using sequences from PLHIV with previously known transmission dates (clinically estimated dates of infection) suggested that the difference between clinically estimated and molecular clock inferred infection dates was smaller for PLHIV infected within MTCs with short branches translated as short time intervals between sampling and infection. On the other hand, this difference was not correlated with the time interval from the last seronegative to the first seropositive tests, suggesting that the discordance between the two infection dates was not due to a larger uncertainty of the clinically estimated dates of infection. The molecular clock inferred age of infection preceded the clinically estimated one for most PLHIV due to the characteristics of HIV transmission. Specifically, the t_MRCA_ of an internal node of viral sequences from two persons (i.e., the source and the recipient) corresponds to the coalescent time of the viral lineages in the source (8). Therefore, the molecular clock inferred dates of infection can precede the clinically estimated dates due to the existing genetic divergence at the time of infection (pre-transmission interval) (8, 9). In our case, only a single date was estimated for both the source and the recipient and thus the accuracy of the infection ages depends also on the serial interval (time between two HIV cases in a transmission chain). This probably explains why the accuracy is higher for PWID, since in this case a large number of transmissions occurred within a short time frame.

To date, molecular markers, such as the low proportion of ambiguity sites in partial HIV-1 sequences, have been shown to provide a reliable marker of recent transmission (5-7). Kouyos *et al* suggested that the proportion of ambiguity sites increased by 0.2% within the first 8 years and a proportion >0.5% is a marker of a non-recent infection (5). Earlier studies have shown that molecular clock analyses using multiple sequences per patient can reliably estimate HIV-1 infection dates (8, 9). More recently, Puller *et al* showed that within patient viral diversity estimated using NGS data can be used for the accurate estimation of infection dates (11). The accuracy of the estimations was improved by increasing the sequence depth and particularly viral diversity in the 3^rd^ codon position increased linearly with time (11). NGS data are superior to the fraction of ambiguous sites or Sanger sequencing (11, 12). Moreover, NGS does not require heterochronous sampling and this provides an additional strength of this method.

In the current study, we show that t_MRCA_ estimations of the infection ages based on Sanger sequencing data can be used reliably as an approximation of HIV infection dates. Our approach has a few limitations, such as that it can be utilized only for sequences from PLHIV infected with MTCs. Our approach is probably less reliable than NGS, however NGS data or heterochronous sequences are largely missing from HIV clinical laboratories. Alternatively, and under specific conditions, widely available HIV-1 Sanger sequencing data can be used for the estimation of HIV infection dates. Finally, if MTCs have been characterized, our study suggests that dating can be used for an on-going epidemic to quickly assess whether a given blood sample represents a recent infection. This is a useful adjunct to LAg for HIV public health interventions in accordance with one of the key strategies to respond quickly to potential HIV outbreaks as part of the Ending HIV plan in the USA.

## Data Availability

We will share our data upon reasonable request.

